## Supplementary Tables for "*Mycobacterium tuberculosis* infection, immune activation, and risk of HIV acquisition"

**Supplemental Table 1: Antibodies, Sources, and Catalogue numbers for Flow Cytometry**

| **Characteristic** | **Antibody** | **Fluorophore** | **Manufacturer** | **Cat#** | **Clone** |
| --- | --- | --- | --- | --- | --- |
| Cell viability | AViD | AmCyan | Thermo Fisher | L34976 |  |
| T-cell lineage | CD3 | ECD | Beckman Coulter | IM2705U | UCHT-1 |
|  | CD4 | Alexa flour700 | eBioscience | 56-0049-42 | RPA-T4 |
|  | CD8 | PerCP Cy 5.5 | BD | 341051 | SK1 |
| Intracellular cytokines | IL2 | PE | BD | 559334 | MQ1-17H12 |
|  | TNF | FITC | Beckman Coulter | 554512 | Mab11 |
|  | IFN-ɣ | APC | BD | 554702 | B27 |
|  | IL-17a | BV421 | Biolegend | 50-403-676 | BL168 |

**Supplementary Table 2: TaqMan Primer-probe Assays included in Correlates of Risk Analyses.** Some primers included in the multiplex Fluidigm assay were unassigned to previously described signatures, and therefore did not contribute to the published analyses

| **Gene** | **Assay Name** | **Catalog #** | **Custom Assay ID** | **Signature** |
| --- | --- | --- | --- | --- |
| KLF2 | KLF2_CUSTOM | 4441114 | ARH6CF4 | Sweeney3 |
| GBP5 | Hs00369472_m1 | 4331182 |  | Sweeney3 |
| KLF2 | Hs00360439_g1 | 4331182 |  | Sweeney3 |
| GAS6 | GA6_NM_000820_10_11* | 4441114 | AR323W3 | Suliman4 |
| GAS6 | GAS6_NM_000820_5_6* | 4441114 | AR47XGZ |  |
| BLK | Hs01017452_m1 | 4331182 |  | Suliman4 |
| CD1C | Hs00957534_g1 | 4351372 |  | Suliman4 |
| SEPT4 | Hs00910208_g1 | 4331182 |  | Suliman4 |
| FCGR1B | Hs02341825_m1 | 4331182 |  | RISK6 |
| GBP2 | Hs00894846_g1 | 4351372 |  | RISK6 |
| SDR39U1 | Hs01016970_g1 | 4351372 |  | RISK6 |
| SERPING1 | Hs00934329_m1 | 4331182 |  | RISK6 |
| TRMT2A | Hs01000041_g1 | 4351372 |  | RISK6 |
| TUBGCP6 | Hs00363509_g1 | 4351372 |  | RISK6 |
| MAP7D3 | Hs00226257_m1 | 4331182 |  | RESPONSE5 |
| RP11-295G20.2 | Hs01373568_m1 | 4351372 |  | RESPONSE5 |
| SMARCD3 | Hs01088251_g1 | 4351372 |  | RESPONSE5 |
| STT3A | Hs00967491_m1 | 4351372 |  | RESPONSE5 |
| UCP2 | Hs01075224_g1 | 4351372 |  | RESPONSE5 |
| GBP1 | Hs00977005_m1 | 4331182 |  | Maertzdorf4 |
| ID3 | Hs00954037_g1 | 4331182 |  | Maertzdorf4 |
| IFITM3 | Hs03057129_s1 | 4331182 |  | Maertzdorf4 |
| P2RY14 | Hs01848195_s1 | 4331182 |  | Maertzdorf4 |
| TMBIM6 | Hs00162661_m1 | 4331182 |  |  |
| CDC42 | Hs03044122_g1 | 4331182 |  |  |
| USF2 | Hs01100994_g1 | 4351372 |  |  |
| ACTR3 | Hs01029159_g1 | 4351372 |  |  |
| ANKRD22 | Hs00944015_m1  Hs00376013_g1 | 4331182 |  |  |
| ARG1 | Hs00968979_m1 | 4331182 |  |  |
| BATF2 | Hs00912737_m1 | 4331182 |  |  |
| C1QB | Hs00608019_m1 | 4331182 |  |  |
| DUSP3 | Hs01115776_m1 | 4331182 |  |  |
| FCGR1A/CD64 | Hs02340031_m1  Hs00174081_m1 | 4331182 |  |  |
| GBP6 | Hs01584201_m1 | 4331182 |  |  |
| GZMA | Hs00989184_m1 | 4331182 |  |  |
| LTF | Hs00158924_m1 | 4331182 |  |  |
| OSBPL10 | Hs00215016_m1 | 4331182 |  |  |
| PRDM1 | Hs00153357_m1 | 4331182 |  |  |
| Rab33A | Hs00191243_m1 | 4331182 |  |  |
| SCARF1 | Hs01092485_g1 | 4351372 |  |  |
| TMCC1 | Hs01037666_s1 | 4331182 |  |  |
| ZNF296 | Hs00377132_m1 | 4331182 |  |  |

*2 GAS6 probes included in the primer set, but only GA6_NM_000820_10_11* included in the Suliman4 signature, as in the methods of the original signature description.^28^

**Supplementary Table 3:** Univariate associations between HIV acquisition risk predictors and Correlates of Risk (CoR) signatures

| **CoR signature** | | **Characteristic level**  **(Mean, SD)** | | | | | | **p-value^a^** | |
| --- | --- | --- | --- | --- | --- | --- | --- | --- | --- |
| **Study Arm** | | **Placebo** | | | **Treatment** | | | |  |
| RISK6^b^ | | -3.33 (1.70) | | | -3.05 (1.69) | | | | 0.24 |
| Suliman4^b^ | | -1.53 (0.92) | | | -1.53 (0.94) | | | | 0.99 |
| Sweeney3 | | -1.82 (1.28) | | | -1.75 (1.47) | | | | 0.85 |
| Maertzdorf4 | | -1.35 (1.31) | | | -1.06 (1.13) | | | | 0.11 |
| RESPONSE5^b^ | | -1.08 (0.51) | | | -1.20 (0.81) | | | | 0.21 |
| **Gender** | | **Male** | | | **Female** | | | |  |
| RISK6^b^ | | -3.31 (1.67) | | | -3.15 (1.71) | | | | 0.73 |
| Suliman4^b^ | | -1.41 (0.95) | | | -1.55 (0.92) | | | | 0.53 |
| Sweeney3 | | -1.97 (1.04) | | | -1.75 (1.44) | | | | 0.53 |
| Maertzdorf4 | | -1.32 (1.25) | | | -1.17 (1.21) | | | | 0.66 |
| RESPONSE5^b^ | | -1.16 (.53) | | | -1.15 (0.72) | | | | 0.90 |
| **Risk Score above or below median** | | **Risk score < 3** | | | **Risk score ≥ 3** | | | |  |
| RISK6^b^ | | -3.05 (1.60) | | | -3.24 (1.76) | | | | 0.53 |
| Suliman4^b^ | | -1.59 (0.96) | | | -1.49 (0.91) | | | | 0.53 |
| Sweeney3 | | -1.75 (1.54) | | | -1.80 (1.29) | | | | 0.88 |
| Maertzdorf4 | | -1.21 (1.12) | | | -1.18 (1.28) | | | | 0.90 |
| RESPONSE5^b^ | | -1.18 (0.70) | | | -1.13 (0.69) | | | | 0.66 |
| **Ad5 titer >18** | | **Ad5 titer >18** | | | **Ad5 titer ≤18** | | | |  |
| RISK6^b^ | | -3.34 (1.78) | | | -3.04 (1.64) | | | | 0.21 |
| Suliman4^b^ | | -1.43 (0.91) | | | -1.60 (0.94) | | | | 0.21 |
| Sweeney3 | | -1.84 (1.38) | | | -1.74 (1.39) | | | | 0.66 |
| Maertzdorf4 | | -1.14 (1.127) | | | -1.24 (1.29) | | | | 0.66 |
| RESPONSE5^b^ | | -1.14 (0.67) | | | -1.16 (0.71) | | | | 0.90 |
| **HSV-2 infection^c^** | | **HSV-2 Negative** | | | **HSV-2 Positive** | | | |  |
| RISK6^b^ | | -3.28 (1.72) | | | -2.92 (1.68) | | | | 0.21 |
| Suliman4^b^ | | -1.52 (0.90) | | | -1.57 (0.95) | | | | 0.82 |
| Sweeney3 | | -1.85 (1.33) | | | -1.57 (1.60) | | | | 0.21 |
| Maertzdorf4 | | -1.18 (1.167) | | | -1.15 (1.31) | | | | 0.90 |
| RESPONSE5^b^ | | -1.09 (0.62) | | | -1.25 (0.87) | | | | 0.21 |
| **Region^d^** | **North America** | | **South America** | **Caribbean** | | **Australia** |  | | |
| RISK6^b^ | -3.41 (1.73) | | -2.52 (1.52) | -3.45 (1.34) | | -3.09 (0.93) | **0.01** | | |
| Suliman4^b^ | -1.54 (0.85) | | -1.51 (1.11) | -1.58 (0.95) | | -1.20 (1.35) | 0.90 | | |
| Sweeney3 | -2.00 (1.16) | | -1.00 (1.77) | -2.08 (1.11) | | -3.01 (0.46) | **0.01** | | |
| Maertzdorf4 | -1.24 (1.21) | | -1.02 (1.26) | -1.10 (0.88) | | -1.84 (1.51) | 0.53 | | |
| RESPONSE5^b^ | -1.03 (0.50) | | -1.46 (0.99) | -1.42 (0.84) | | -0.92 (0.22) | **0.01** | | |

^a^ Test statistics were calculated to compare across variable levels for each CoR score. Two-sided t-tests were used for binary variables and oneway ANOVA was used for categorical variables. P values adjusted for false discovery with BH correction. BH adjusted p values <0.05 are bolded.

^b^Log transformed

^c^Only performed in men, so this analysis restricted to male participants

^d^Regions include the following countries. North America: USA, Canada; South America: Peru, Brazil; Caribbean: Haiti, Dominican Republic; Australia.

**Supplementary Table 4:** Primary outcomes reported in a subset of the study in which samples were collected a) ≤365 days before HIV diagnosis (n=282) and b) 6 months or less (≤183 days) before HIV diagnosis (n=159). The results below report on the association of all primary variables of interest (LTBI status, *Mtb*-specific CD4 T-cell activation, Correlate of Risk signatures) and HIV acquisition within this sample subset. Associations with a ≥10% change from the primary analysis are bolded.

4A. ≤365 days before HIV diagnosis

|  | Univariate regression estimates | Multivariate regression estimates |
| --- | --- | --- |
|  | OR^a^ (95% CI) | aOR^a,b^ (95% CI) |
| LTBI Positive | 0.87 (0.46, 1.65) | 0.91 (0.46, 1.79) |
| PFS^c^ | 0.99 (0.77, 1.28) | 1.00 (0.77, 1.31) |
| FS^c^ | 1.00 (0.78, 1.27) | 1.00 (0.78, 1.30) |
| RISK6^c,d^ | 1.10 (0.83, 1.44) | 1.28 (0.94, 1.75) |
| Suliman4 ^c,d^ | 0.88 (0.68, 1.13) | 0.90 (0.69, 1.19) |
| Sweeney3 ^c^ | 1.47 (1.06, 2.04) | **1.56 (1.10, 2.21)** |
| Maertzdorf4 ^c^ | 0.96 (0.74, 1.25) | 0.97 (0.74, 1.28) |
| RESPONSE5 ^c,d^ | 0.76 (0.55, 1.06) | 0.68 (0.47, 0.97) |

4B. ≤183 days before HIV diagnosis

|  | Univariate regression estimates | Multivariate regression estimates |
| --- | --- | --- |
|  | OR^a^ (95% CI) | aOR^a,b^ (95% CI) |
| LTBI Positive | **1.07 (0.45, 2.51)** | **1.09 (0.41, 2.90)** |
| PFS^c^ | **1.19 (0.82, 1.72)** | **1.29 (0.85, 1.95)** |
| FS^c^ | **1.17 (0.83, 1.64)** | **1.27 (0.87, 1.87)** |
| RISK6^c,d^ | **1.20 (0.81, 1.77)** | 1.26 (0.81, 1.96) |
| Suliman4 ^c,d^ | 0.97 (0.70, 1.34) | 0.92 (0.65, 1.31) |
| Sweeney3 ^c^ | **1.62 (1.00, 2.61)** | **1.59 (0.93, 2.70)** |
| Maertzdorf4 ^c^ | **1.10 (0.76, 1.59)** | **1.00 (0.69, 1.46)** |
| RESPONSE5 ^c,d^ | **0.68 (0.40, 1.17)** | **0.65 (0.36, 1.16)** |

Note that although most of the estimates in Table 3 changed by >10% the small sample size reduced power, such that there was likely instability in the point estimate, and loss of statistical significance (when present in the full dataset), such that no new meaningful associations arose from the time-limited sensitivity analyses. Some sensitivity analyses used less than the total number of participants meeting the time criteria due sample viability or missingness, as described in the primary analysis (maximum not included from total: n=11 for LTBI, FS, PFS and n=26 for all CoR scores).

^a^All models used conditional logistic regression accounting for matching variables (treatment arm, country)

^b^Models were fully fit with all variables that were associated with case control status: gender, risk score, and age.

^c^ORs are reported for 1 standard deviation change.

^d^Log transformed

| CoR Score^c^ | LTBI Negative | LTBI Positive |
| --- | --- | --- |
|  | aOR^a^ (95% CI) | aOR^a^ (95% CI) |
| RISK6^b^ | 1.24 (0.97, 1.59) | 0.58 (0.34, 0.99) |
| Suliman4^b^ | **0.77 (0.61, 0.97)** | 1.15 (0.69, 1.91) |
| Sweeney3 | **1.40 (1.08, 1.81)** | 0.81 (0.48, 1.37) |
| Maertzdorf4 | 0.96 (0.76, 1.22) | **0.29 (0.14, 0.64)** |
| RESPONSE5^b^ | **0.76 (0.61, 0.96)** | 0.90 (0.51, 1.56) |

### Supplementary Table 5: Associations between each Correlates of Risk (COR) signature and HIV acquisition, adjusted analyses stratified by LTBI status

*Significant BH adjusted p-values (α=0.05) are presented in bold type.

^a^OR and adjusted OR (aOR) are reported for 1 standard deviation change.

^b^Variables log transformed.

^c^All CoR scores were modeled using logistic regression controlling for matching variables (treatment arm, country) as well as all variables that were associated with case control status: gender, risk score, and age. Conditional logistic regression not used in stratified analysis as matched cases and controls could have differing LTBI status, resulting in exclusion of entire LTBI-discordant matched sets from the models.

**Supplementary Table 6: Univariate associations between Correlates of Risk Scores (CoR) and Latent Tuberculosis Infection (LTBI) status**

| CoR Score | Univariate regression estimates for LTBI+ |
| --- | --- |
|  | OR^a^ (95% CI) |
| RISK6^b^ | 0.85 (0.63, 1.14) |
| Suliman4^b^ | 1.03 (0.77, 1.38) |
| Sweeney3 | 0.99 (0.75, 1.31) |
| Maertzdorf4 | 1.00 (0.72, 1.41) |
| RESPONSE5^b^ | 1.19 (0.88, 1.62) |

^a^ORs are reported for 1 standard deviation change.

^b^Log transformed
